# Concentration of the cellular material in the nasopharyngeal swabs increases the clinical sensitivity of SARS-CoV2 RT-PCR

**DOI:** 10.1101/2020.10.31.20218958

**Authors:** Priya Kannian, Pasuvaraj Mahanathi, Veeraraghavan Ashwini

## Abstract

Severe acute respiratory syndrome - coronavirus 2 (SARS-CoV2) is detected by a highly sensitive molecular method, reverse transcriptase-polymerase chain reaction (RT-PCR) from nasopharyngeal swab (NPS) samples collected in 2-3ml of viral transport medium (VTM). Unlike body fluids, NPS samples are undermined by high variability in the amount of cells that get suspended into the VTM. Hence, the cell density used for RNA extraction becomes an important analytical variable that contributes to the overall sensitivity of the RT-PCR. In this study, we compared the sensitivity of SARS-CoV2 RT-PCR in 50 NPS samples collected from in-patients of the COVID wards using the concentration and direct methods. The concentration method detected the viral RNA in all 50 samples, while the direct method was positive in only 41 (82%) samples (p=0.003). Additionally, the Ct values were lower in the direct method compared to concentration method among the 41 positive samples (p=0.03 for *N gene* and p=0.04 for *RdRp gene*). The mean CV% was also ≥10%. Thus, the concentration of the cells prior to RNA extraction drastically improves the sensitivity of detection of SARS-CoV2 in NPS samples.

---

Severe acute respiratory syndrome - coronavirus 2 (SARS-CoV2) is a single stranded RNA virus that infects the epithelial cells of the lungs and the respiratory tract. The current gold standard for the detection of SARS-CoV2 is reverse transcriptase-polymerase chain reaction (RT-PCR). RT-PCR is a molecular method with a very high analytical sensitivity that can detect even a single copy of RNA per reaction due to the million-fold amplification. However, the clinical sensitivity of RT-PCR to detect SARS-CoV2 has been determined to be about 70% (Wang *et al*., 2020). Clinical sensitivity is always 10-100 folds lower than the analytical sensitivity because of the presence of many chemical mediators in the clinical samples that could potentially inhibit the PCR. Additionally, a number of other pre-analytical and analytical factors contribute to the reduced clinical sensitivity. A few of these pre-analytical issues have already been addressed including type of sample, the medium used for collection and transport of nasopharyngeal swab (NPS), and storage time and temperature (Basso *et al*., 2020).

NPS has been widely used as the most reliable specimen for the detection of SARS-CoV2. NPS upon collection is immediately placed into 2-3ml of viral transport medium (VTM). Any specimen collected on a swab has an inherent limitation in the retrieval of the cells during processing. The best possible retrieval method is to vortex the swab in the VTM before removing it. The volume of VTM containing the virus-infected cells used for RNA extraction plays an important role in the sensitivity of the detection of the virus, as it is directly proportional to the cell density. A number of studies have used varying volumes of NPS for the detection of respiratory viruses (table 1). In contrast, body fluids like plasma, pleural fluid, nasopharyngeal aspirate, etc contain a more homogenous mixture of the cellular material. Thus, a NPS sample in VTM is different from body fluids, but similar to a tissue culture sample, where the cells are suspended in a conditioned medium. Usually, for DNA/RNA extraction from tissue cultures, the cells will be pelleted down by centrifugation (manufacturer’s instructions in DNeasy / RNeasy blood and tissue kits, Qiagen, USA). The medium will be aspirated out and the cell pellet will be lysed in lysis buffer. This method facilitates the concentration of the cells thereby increasing the sensitivity of detection. Therefore, in the current study we evaluated the difference between the concentrated sample and the direct sample of NPS in the detection of SARS-CoV2 RNA.

**Table 1:** Variable volumes used for RNA extraction for the detection of respiratory viruses from nasopharyngeal swab samples collected in viral transport medium by many reported studies

| S. No. | Article | Sample volume |
| --- | --- | --- |
| 1 | Chandler *et al* 2012 | 250µl |
| 2 | Griesemer *et al* 2013 | 250µl |
| 3 | Jeong *et al* 2014 | 300µl |
| 4 | Kim *et al* 2014 | 150µl |
| 5 | Piralla *et al* 2018 | 500µl |
| 6 | Basso *et al* 2020 | 300µl |
| 7 | CDC 2020 | 140µl |

At VHS Laboratory Services, VHS Hospital, Chennai, India NPS samples are routinely processed by the concentration method (Method I) for the detection of SARS-CoV2 RNA by RT-PCR. The NPS is placed in 3ml of VTM and transported immediately to the laboratory in cold chain. The tube is vortexed gently for 15 seconds and then the swab is removed. Two hundred microlitres of the sample was stored for method II. The samples were centrifuged at 3500 rpm for 10 minutes. Supernatants were discarded leaving 750µl of VTM, in which the cell pellets were resuspended. From here 200µl of sample was lysed with 560µl of RNA lysis buffer (QIAamp viral RNA mini kit, Qiagen, USA). After manual lysis, the samples were loaded onto the QIA Cube Connect (Qiagen, USA) for the automated extraction of RNA. Finally, the RNA was eluted in 60µl of AVE buffer (provided in the kit). For RT-PCR, 5µl of the eluted RNA was added to 15µl of RT-PCR mastermix (Labgun COVID-19 assay plus, Labgenomics, Korea; manufacturer validated limit of detection is five copies per reaction). The reactions were amplified using Lightcycler 96 (Roche, USA) as per the manufacturer’s instructions.

For the retrospective comparative method study, 50 NPS positive samples from patients admitted in the COVID wards were included in an anonymous delinked manner. As this a qualitative RT-PCR with 45 amplification cycles, cycle threshold (Ct) value of 25 was used as a cut-off to stratify samples with a high / low viral burden. Twenty five samples with a high Ct value (≥ 25) and 25 samples with a low Ct value (≤ 24) were selected. For the direct sampling method (method II), 200µl of the stored NPS sample was mixed with 560µl of RNA lysis buffer. These lysates were processed as per the protocol given in method I.

For statistical analyses, mean and standard deviation were calculated using Microsoft Excel. Coefficient of variation was calculated using the formula, CV% = SD/ Mean multiplied by 100. For t-test of two independent means calculation, the online calculator was used.

Of the 50 NPS samples, only 41 (82%) were positive by both the methods. Nine (18%) samples belonging to the low viral burden group (>25 Ct) were negative by the direct method (method II), indicating lower detection rate. This difference between the two methods was statistically significant (p=0.003; Fisher Exact Probability test). Since this is a qualitative RT-PCR, the sensitivity of the assay was determined by comparing the Ct values. A lower Ct value indicates higher copy numbers or higher viral burden. In Figure 1, the Ct values of all 41 positive samples for both the *N gene* and *RdRp gene* are shown for both methods I and II. The mean Ct value of method I (*N gene* mean Ct is 23 and *RdRp gene* mean Ct is 25) is 2-3 cycles lower than that of method II (*N gene* mean Ct is 26 and *RdRp gene* mean Ct is 27), indicating higher sensitivity of detection by method I. This difference was statistically significant (Figure 1). The CV% was more than or equal to 10% for both the *N gene* and *RdRp gene* (table 2). These findings together indicate that the sensitivity of method I (concentration method) to detect the SARS-CoV2 RNA is superior to that of method II (direct method).

**Table 2:** Coefficient of variation between the concentration and direct methods of sampling from nasopharyngeal swabs (n=41) for the detection of SARS-CoV2 by RT-PCR

| S. No. | Gene name | CV% - Mean | CV% - Range | CV - <5% | CV - >5% |
| --- | --- | --- | --- | --- | --- |
|  |  |  |  | n (%) | n (%) |
| 1 | IC | 6.73 | 0.2-27.8 | 29 (70.7) | 12 (29.3) |
| 2 | N | 10.99 | 0.5-39.8 | 8 (19.5) | 33 (80.5) |
| 3 | RdRp | 9.75 | 0.9-36.9 | 5 (12.2) | 36 (87.8) |

CV – coefficient of variation
IC – internal control gene; N – nucleocapsid gene; RdRp – RNA dependent RNA polymerase gene
n – number of samples that had <5% or >5% CV
% - n/41 multiplied by 100

**Figure 1:**
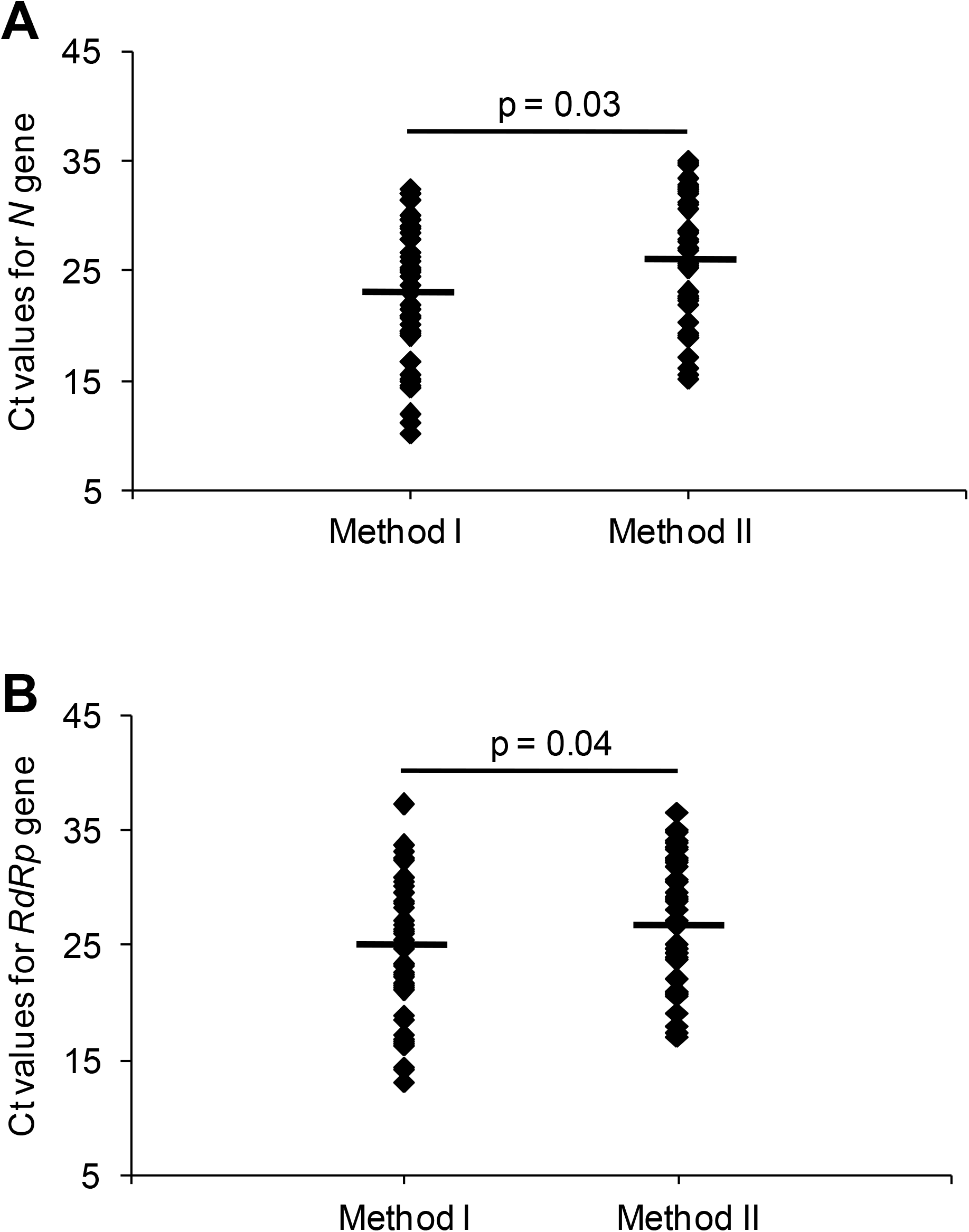
Comparative Ct values for *N gene* and *RdRp gene* by RT-PCR using two sampling methods. X axis denotes the two methods: Method I - concentration method and Method II - direct method. Y-axis denotes the Ct values. The diamond represents the 41 nasopharyngeal swab samples. The short black bar represents the mean. The p value was calculated by independent t-test. **A**. *N gene*. **B**. *RdRp gene*.

The concentration method has its own caveat. While concentrating the cellular material, the PCR inhibitors that are present in the sample will also get concentrated simultaneously, thereby increasing the potential possibility of false negatives. This issue was addressed by analyzing the detection of the internal control (IC) gene. The CV% of the IC gene was less than 10% (6.73%; Table 2). Since the range of the CV% was wide, we also analysed the percentage of samples that had a low CV% (<5%) and a high CV% (>5%). Majority of the samples had a low CV% for IC gene, but a high CV% for the SARS-CoV2 genes (Table 2). CV% is a clear indicator of the variation between the two results. The low CV% among the Ct values for IC gene indicates that there is no significant variation the detection of the IC gene by both the methods. The IC gene is a positive extraction control that is used to determine the proper extraction of the RNA with negligible or minimal amount of PCR inhibitors. Thus, the data clearly rules out the possibility of concentration of the PCR inhibitors in method I.

In conclusion, our study clearly indicates the value of concentrating the cells in the NPS samples in terms of increased sensitivity in the detection of SARS-CoV2 RNA by RT-PCR. This study addresses yet another analytical issue that could reduce false negativity, which is crucial in preventing transmission of SARS-CoV2.

## Data Availability

Not applicable

## Acknowledgements

This work was funded by Intramural Research Funds of The Voluntary Health Services, Chennai, India.

